# Exclusion of the commonest subtype of B-cell leukemia in children acquiring EBV infection in early life

**DOI:** 10.64898/2026.07.20.26358339

**Authors:** Jutatip Panaampon, Zhe Wang, Il-Kyu Choi, Jiankun Guan, Catherine Seaman, Shanna Richard, Victoria Koch, Marian H. Harris, Yael Flamand, Jerome Ritz, Michael E. Scheurer, Lynda M. Vrooman, Andrew E. Place, Melissa Burns, Lewis B. Silverman, Yana Pikman, Baochun Zhang

**Affiliations:** Department of Medical Oncology, Dana-Farber Cancer Institute, Boston, MA 02215, USA; Department of Medicine, Harvard Medical School, Boston, MA, USA; Department of Pediatric Oncology, Dana-Farber Cancer Institute, Boston, MA; Department of Pathology, Boston Children’s Hospital, Boston, MA; Department of Data Science, Dana-Farber Cancer Institute, Boston, MA; Department of Pediatrics, Emory University School of Medicine, and Children’s Healthcare of Atlanta, Atlanta, GA 30322, USA; Department of Pediatric Oncology, Dana-Farber Cancer Institute; Division of Hematology/Oncology, Boston Children’s Hospital; Harvard Medical School, Boston, MA; Division of Pediatric Hematology, Oncology and Stem Cell Transplantation, Columbia University Irving Medical Center, Vagelos College of Physicians and Surgeons, New York, NY; Department of Cancer Immunology and Virology, Dana-Farber Cancer Institute, Boston, MA 02215, USA; Department of New Biology and New Biology Research Center (NBRC), Daegu Gyeongbuk Institute of Science and Technology (DGIST), Daegu 42988, Republic of Korea

## Abstract

In developed countries, the rate of childhood B-cell acute lymphoblastic leukemia (B-ALL), the most common pediatric cancer with a peak incidence at 2–5 years of age, has been rising for several decades. Epidemiological studies suggest that reduced exposure to common infections in early life increases the risk of B-ALL. However, no specific infection capable of protecting against such cancer has been identified. One of the most prevalent infectious agents in humans is Epstein-Barr virus (EBV), a B-cell tropic tumor virus that infects ~95% of the global population by adult age. Paradoxically, recent studies reveal that EBV, through its signaling protein LMP1, elicits potent cytotoxic CD4^+^ and CD8^+^ T cell responses against a wide range of tumor-associated antigens (TAAs), which can recognize and attack EBV-unrelated cancer cells via shared TAAs. In developed countries, primary EBV infection is often delayed from early childhood into adolescence or young adulthood. Taken together, we hypothesized that EBV (LMP1)-induced TAA-specific T cells may help protect against some childhood B-ALL by targeting shared TAAs. If so, lack of EBV infection in early life may contribute to the rise of childhood B-ALL seen in developed countries. In this work, EBV serology assessment in pediatric B-ALL patients revealed strong exclusion of the commonest high hyperdiploid (HHD) subtype of B-ALL in children having recent primary EBV infection. Our mouse model studies demonstrated that LMP1-induced T cell immunity can eradicate some B-ALL–like leukemias via shared TAAs during the effector phase. These findings support the notion that EBV-induced anti-tumor immunity may help protect against some childhood B-ALL.

## INTRODUCTION

The rate of childhood cancer has been rising at ~0.7% per year for several decades in the US and other developed countries, in sharp contrast to the roughly stable overall rate of adult cancer^1^ (Fig. S1A). The rise involves many types of childhood cancer, including B-cell acute lymphoblastic leukemia (B-ALL), which is the most common pediatric cancer with a peak incidence at 2–5 years of age^2^ (Fig. S1B; Fig. S2). Epidemiological studies suggest that reduced exposure to common infections in early life increases the risk of B-ALL in developed countries^2,3^. However, thus far, no specific infection capable of protecting against cancer has been identified, and any putative underlying mechanisms remain a matter of speculation and debate^2,4-6^.

One of the most common infectious agents in humans is Epstein-Barr virus (EBV), a B-cell tropic tumor virus that infects ~95% of the global population by adult age. Paradoxically, recent studies^7^ reveal that EBV, through its signaling protein LMP1, elicits potent anti-tumor immunity. This includes cytotoxic CD4^+^ and CD8^+^ T cell responses against a wide range of tumor-associated antigens (TAAs)^7^, which can recognize and attack EBV-unrelated cancer cells via shared TAAs^8,9.^ In developed countries, primary EBV infection is often delayed from early childhood into adolescence or young adulthood^10^. We hypothesized^1^ that EBV (LMP1)-induced TAA-specific T cells may help protect against some childhood B-ALL by targeting shared TAAs. If so, lack of EBV infection (and its induced anti-tumor immunity) in early life may contribute to the rise in the rate of childhood B-ALL seen in developed countries.

To address the hypothesis, in this work we tested the potential of LMP1-induced T cells in protection against B-ALL–like leukemias in mouse models, and conducted EBV serology study using plasma samples from pediatric patients with B-ALL enrolled on the Dana-Farber Cancer Institute (DFCI) ALL Consortium Studies 05-001 (NCT00400946)^11,12^ and 11-001 (NCT01574274). The results from these studies together suggest that primary EBV infection in early years, particularly at ages 2–3, may prevent the commonest high hyperdiploid (HHD) subtype of childhood B-ALL in most cases.

## RESULTS

### Significant negative association between EBV seropositivity and the commonest subtype of B-ALL in children at early ages

In seeking evidence that primary EBV infection may protect/prevent some B-ALL in childhood, we first attempted to test a correlation between EBV infection and pediatric B-ALL (pB-ALL), by testing the rate of EBV prevalence in pB-ALL patients vs healthy controls. However, given the impact of a number of key factors influencing EBV exposure, such as race/ethnicity and social/economic status^13,14^, which are usually poorly annotated on banked healthy control and patient plasma samples, this turned out to be an impractical approach.

We then turned to testing for an association between EBV infection and subtypes of pB-ALL, focusing on the two most common subtypes, high hyperdiploid (HHD; 51-65 chromosomes) and *ETV6::RUNX1* (*E/R*)^15^ due to sample availability. We had patients enrolled on DFCI ALL Consortium clinical trials DFCI 05-001 (2005-2011, n=800), 11-001 (2012-2015, n=240), and 16-001 (2017-2022, n=560). The number and frequency of patients enrolled per age and genetic subtype mirrored the pattern of B-ALL incidence in the US (Figs. 1A–B, Fig. S2) and UK^16^. We focused on patients with HHD and *E/R* subtypes of B-ALL, which account for 26% and 20% of the full cohort, and 35% and 29% of the disease at ages 2–5 years old, respectively, with this age having the highest incidence of B-ALL. All other genetic subtypes were grouped together into “Others” category.

**Figure 1.**
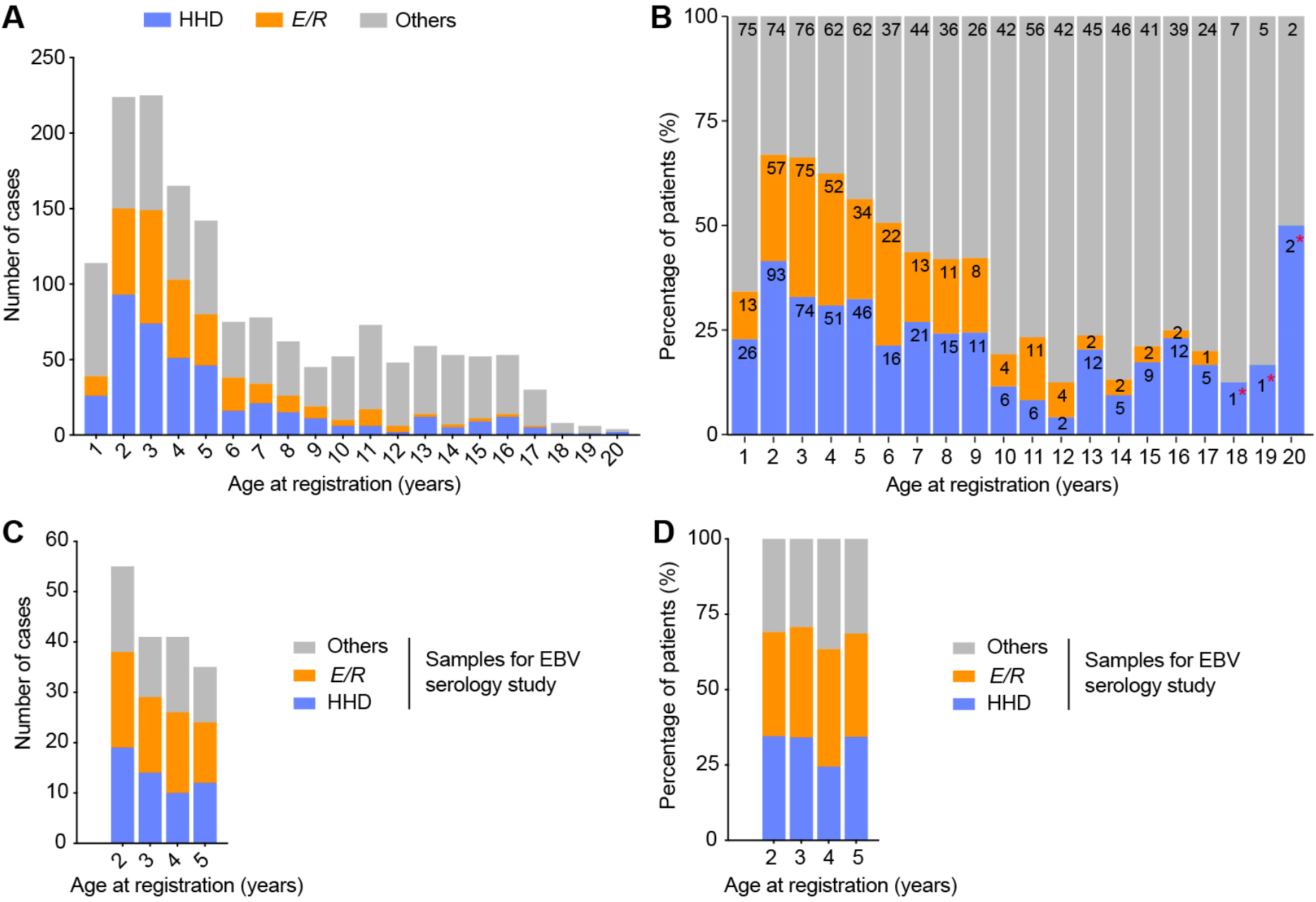
Age distribution of HHD, *E /R*, and Others subtypes of B-ALL cases pooled from DFCI/BCH cohorts, and those used in this study. **A**, Overview of the number of cases of HHD, *E /R*, and Others subtypes of B-ALL at each indicated age. **B**, Percentages of HHD, *E /R*, and Others subtypes of B-ALL (color keys as in A) at each indicated age; numbers of cases are also shown in the colored bars. *Given the small sizes of these cohorts, these percentages are not considered precise. In A-B, cases were pooled from DFCI/BCH cohorts 05-001, 11-001 and 16-001. **C-D**, Numbers (C) and fractions (D) of samples of the indicated subtypes of B-ALL at each indicated age included in the EBV serology study; samples pulled from the cohorts 05-001 and 11-001.

We tested plasma samples from patients with B-ALL diagnosed at ages 2–5 for EBV serology. The sample numbers of the HHD, *E/R*, and “Others” categories of B-ALL were in accord with their respective ratios at each of these ages (Figs. 1C–D). There was no other selection criterion, and the samples were selected randomly from those banked and available from the DFCI 05-001 and DFCI 11-001 studies. In total, 179 samples were subjected to EBV serostatus assessment. Current technologies can determine acute and past EBV infection based on the kinetics of EBV-specific antibody responses^17^. The ‘acute infection’ phase can be detected by IgM antibody positivity to EBV viral capsid antigen (VCA), which has a short 1–1.5 months of detection window^18^. The ‘past infection’ phase lasts for the duration of life and can be detected by VCA IgG positivity. There is currently no way to pinpoint how long it has been since the initial infection. We performed EBV antibody testing using ELISA kits^7,13^,14, resulting in 172 samples with unambiguous readout. In these samples, no acute EBV infection was detected, while past EBV infection was found in 82/172 (47.7%) samples (Table 1).

**Table 1.** EBV infection status of B-ALL patients determined by antibody test in plasma samples collected at the time of leukemia diagnosis.

|  |  |  |  |  |
| --- | --- | --- | --- | --- |
|  | Subject No. | No infection<br>(VCA IgM<sup>-</sup>, VCA IgG<sup>-</sup>)<br>Subject No. (%) | Acute infection<br>(VCA IgM<sup>+</sup>, VCA IgG<sup>+/-</sup>)<br>Subject No. (%) | Past infection<br>(VCA IgM<sup>-</sup>, VCA IgG<sup>+</sup>)<br>Subject No. (%) |
| Total | 172 | 90 (52.3%) | 0 | 82 (47.7%) |
| Age (year) |  |  |  |  |
| 2 | 55 | 30 (54.5%) | 0 | 25 (45.5%) |
| 3 | 41 | 24 (58.5%) | 0 | 17 (41.5%) |
| 4 | 41 | 18 (43.9%) | 0 | 23 (56.1%) |
| 5 | 35 | 18 (51.4%) | 0 | 17 (48.6%) |

Detailed analysis revealed a statistically significant inverse correlation between HHD B-ALL and EBV seropositivity at ages 2–3; this negative correlation was not seen at ages 4–5 (Figs. 2A-B). In contrast, the *E/R* and “Others” categories exhibited no correlation with EBV seropositivity at ages 2–5. Thus, this serology study reveals a strong exclusion of HHD B-ALL in children who had evidence of an EBV infection prior to the B-ALL diagnosis at ages 2–3 years old, with an odds ratio of 0.22 (95% confidence interval: 0.08-0.57) comparing EBV-seropositive versus - seronegative individuals.

**Figure 2.**
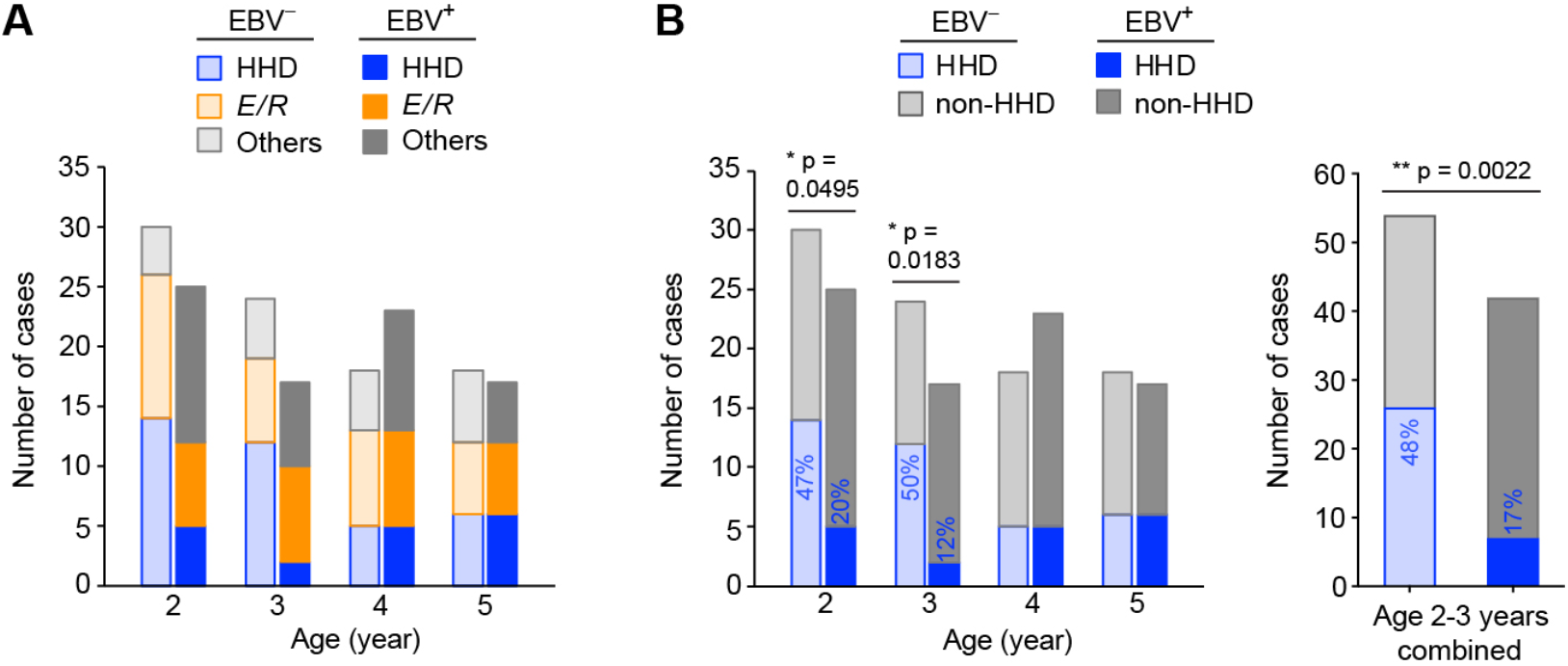
Negative association of EBV seropositivity with HHD B-ALL at ages 2–3. **A**, Distribution of the most prevalent HHD subtype, the second prevalent *E /R* subtype, and Others subtypes of B-ALL in relationship with EBV serostatus at the indicated ages. **B**, Analysis of correlation of the HHD subtype of B-ALL with EBV serostatus at the indicated ages. Statistical analysis by Fisher’s exact test.

### LMP1-induced T cells at effector phase can protect against B-ALL–like leukemia in mice

We used a mouse model to study the mechanism of immune surveillance against development of leukemia in children. Our previous studies revealed a key role of LMP1 in EBV-induced immunity^19^ and characterized an inducible LMP1-expressing *CD19-cre*^*ERT2*^*;LMP1*^*flSTOP*^ (*ERT2-CL*) mouse model. In these mice, a single dose (3-4 mg) of tamoxifen (Tmx; administered by intragastric gavage) induces LMP1 expression in a small fraction of B cells, which quickly expand over a few days and provoke TAA-specific CD4^+^ and CD8^+^ cytotoxic T lymphocyte (CTL) responses. TAA-specific CD8^+^ and CD4^+^ CTLs peak around day 5, and lead to clearance of LMP1^+^ B cells by day 8. Afterward, TAA-specific T cells contract to baseline levels and form long-term memory^7,20^. We also demonstrated that the T cells primed by LMP1^+^ B cells display MHC-restricted recognition and killing of several LMP1-negative B-lymphoma cell lines *in vitro*, indicative of their ability to target shared TAAs^7^.

Using *ERT2-CL* and control *CD19-cre*^*ERT2*^ (*ERT2-C*) mice, we explored whether the LMP1-induced immunity can protect against mouse B-ALL *in vivo*; for the latter, we used syngeneic cell lines (all expressing pre-B cell markers) established from the aggressive mouse B-ALL model driven by an *Eμ-Myc* transgene^21^ (see Methods). We engrafted the mouse B-ALL cells into *ERT2-CL* and control *ERT2-C* mice, and 3 days later treated the mice with Tmx (see schema in Fig. 3A) to test whether the TAA-specific effector T cells induced by LMP1^+^ B cells have protective effect. Of the four leukemia lines tested (L10, L12-33, L12-3, and L14), we observed complete protection against L10 in all *ERT2-CL* mice and against L12-33 in most *ERT2-CL* mice, but only transient protection against L12-3 and L14 in *ERT2-CL* mice (Fig. 3B). These findings indicate the heterogenous nature of the leukemia lines. Interestingly, when analyzed for leukemia burden in peripheral blood at day 21, all *ERT2-CL* mice inoculated with the four leukemia lines (except one mouse receiving L12-33) had either no detectable or “minimal residual” leukemic cells (Fig. 3C); the latter would be particularly true for *ERT2-CL* mice receiving L12-3 and L14 leukemic cells, given their eventual succumbing to leukemia relapse (Fig. 3B and Fig. S3).

**Figure 3.**
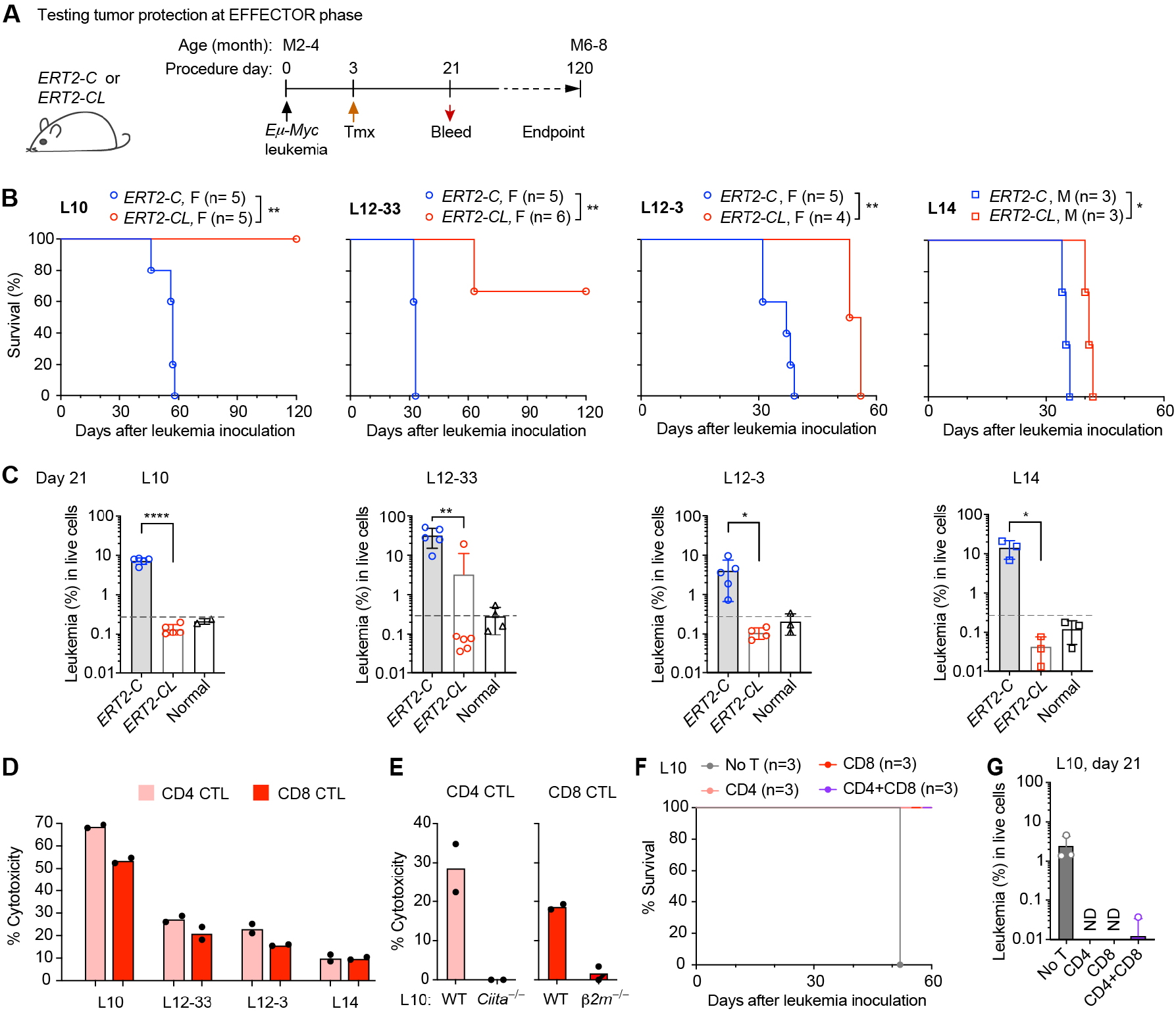
Immunity induced by LMP1^+^ B cells, at effector phase, can protect against leukemia. **A**, Schema of the experimental protocol to test tumor protection at effector phase. Tmx, tamoxifen. **B**, Survival of the mice transplanted with the indicated mouse leukemia lines and treated as in A. F, female; M, male. Survival curves were compared by the log-rank (Mantel-Cox) test. **C**, Leukemia (CD19^+^B220^low^IgM^−^c-Kit^−^) burden assessed by FACS at day 21 in peripheral blood of *ERT2-C* and *ERT2-CL* mice treated as in A. Normal, untreated wild-type mice as control to indicate background staining. Data are presented as mean ± SD. Leukemia burden was compared by unpaired two-tailed Student’s *t* test. **D**, Cytotoxicity of CD4^+^ and CD8^+^ CTLs, isolated from *ERT2-CL* mice carrying a *Foxp3*^*DTR/GFP*^ allele (to allow excluding CD4^+^ T_reg_) 6 days after Tmx treatment, against the indicated leukemia cells at an effector-to-target (E:T) cell ratio of 50:1. Data represent three independent experiments, in each of which T cells were isolated from one mouse and tested against the four leukemia lines. **E**, Cytotoxicity of CD4 and CD8 CTLs, prepared as described in D, against wild-type (WT) L10, or MHC-II^−^ (*Ciita*^*–/–*^) or MHC-I^−^ (*b2m*^*–/–*^) subline generated by CRISPR-Cas9. **F**, Adoptive CD4 and/or CD8 CTLs in control of the L10 leukemia in *Rag2*^*–/–*^*γc*^*–/–*^ hosts. At day 3 after leukemia inoculation, mice received adoptive transfer of T cells isolated from *ERT2-CL* mice and then were monitored for survival. **G**, Leukemia burden assessed by FACS at day 21 in peripheral blood of mice treated as in F.

By *in vitro* killing assay, we found that CD4^+^ and CD8^+^ effector T cells isolated from *ERT2-CL* mice at day 6 post-Tmx exhibited cytotoxicity against the four leukemia lines, with the potency against L10 > L12-33 > L12-3 > L14 (Fig. 3D), a pattern correlating well with their efficacy in controlling these leukemias *in vivo* (Fig. 3B). Additional experiments with L10 cells demonstrated that CD4^+^ and CD8^+^ T cells killed the leukemia cells in MHC-restricted manner (Fig. 3E), indicating that they recognized and killed the latter via shared TAAs presented on MHCs. Furthermore, by adoptive transfer experiments in *Rag2*^*–/–*^*γc*^*–/–*^ hosts, we showed that these T cells were sufficient to eliminate the L10 leukemic cells *in vivo* (Fig. 3F-G)

Collectively, these data indicate that LMP1-induced CD8^+^ and CD4^+^ CTLs, at effector phase, can protect against B-cell leukemias by targeting shared TAAs, and this potentially leads to complete elimination of certain leukemias (such as L10 and L12-33) while only transient control of other leukemias (such as L12-3 and L14), presumably due to their differential immunogenicity.

### No leukemia protection detectable at T-cell memory phase

We next treated *ERT2-CL* and control mice with Tmx, and 4 months later gave leukemia challenge (see schema in Fig. 4A) to test whether the TAA-specific memory T cells induced by LMP1^+^ B cells also have protective effect. However, no protection against any of the four leukemia lines was detected (Fig. 4B and data not shown).

**Figure 4.**
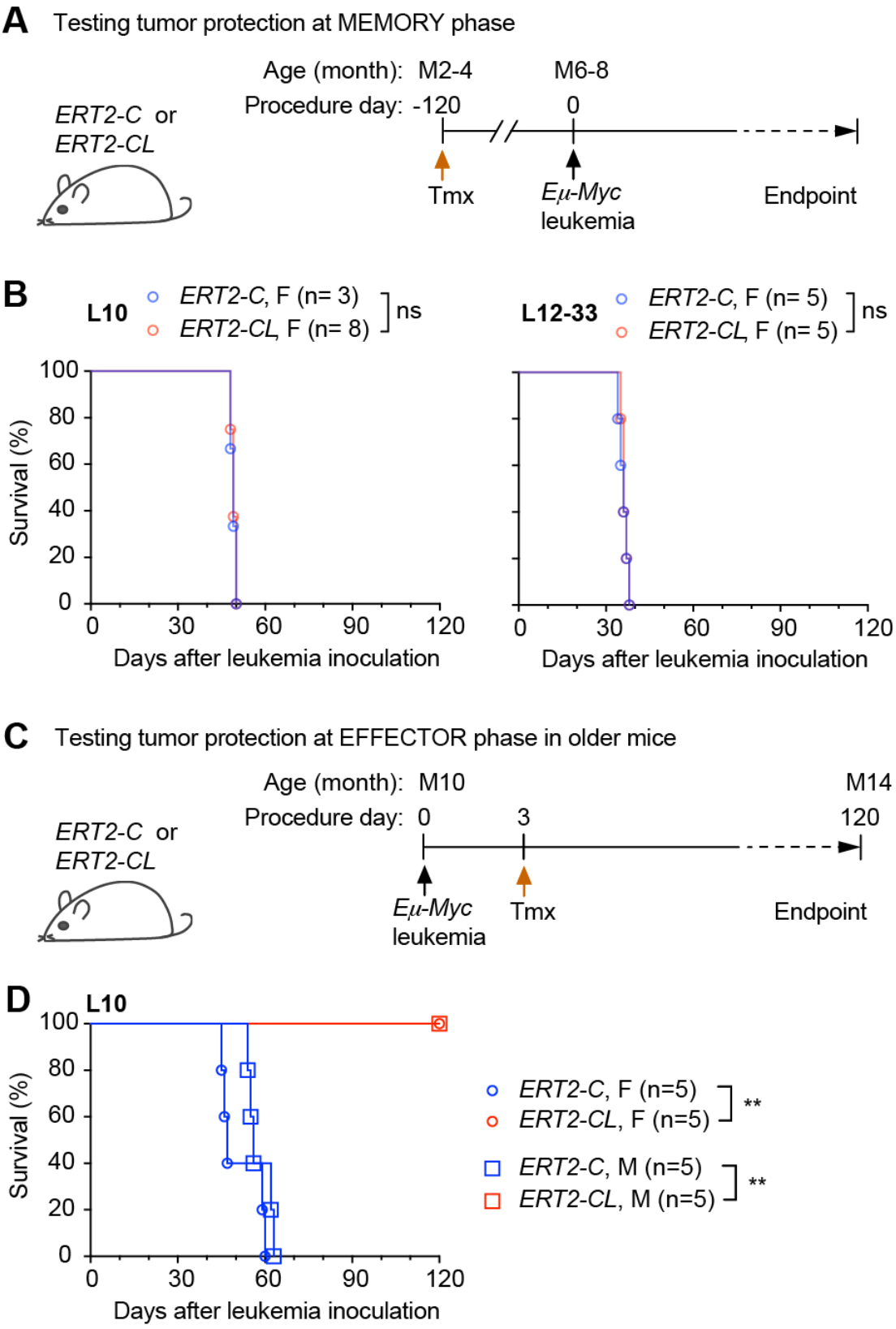
Immunity induced by LMP1^+^ B cells loses leukemia protection at memory phase. **A**, Schema of the experimental protocol to test tumor protection at memory phase. **B**, Survival of the mice transplanted with the indicated mouse leukemia lines and treated as in A. **C**, Schema of the experimental protocol to test tumor protection at effector phase in older mice. **D**, Survival of the mice transplanted with the indicated mouse leukemia line and treated as in C. Mouse survival curves were compared by the log-rank (Mantel-Cox) test.

To confirm that the lack of leukemia protection in this setting was truly due to the incapability of memory T cells, rather than other age-related factors, we performed an additional set of *in vivo* experiments. We engrafted the L10 cells into 10-month-old *ERT2-CL* and control *ERT2-C* mice, and 3 days later treated the mice with Tmx (see schema in Fig. 4C) to test whether the TAA-specific effector T cells induced by LMP1^+^ B cells in older mice have a protective effect against the leukemia. Indeed, the TAA-specific effector T cells in these older mice were similarly potent in leukemia protection as their counterparts in younger mice (Fig. 4D).

Taken together, the results from mouse modelling suggest that TAA-specific T cells have potent tumor protective potential at the effector phase, but they lose such capacity at the memory phase. This informs that the strongest tumor protection by EBV-induced T cells in humans would be around primary EBV infection, which often occurs in early childhood, also the peak time of pediatric B-ALL.

## DISCUSSION

Our results suggest that primary EBV infection in early years, prior to 2–3 years old, can largely preclude the development of HHD B-ALL, the most common subtype of B-ALL in that age group. In mouse studies, we modeled leukemia protection induced by EBV LMP1 that may help interpret this clinical observation. Our findings suggest that TAA-specific T cells have the most potent tumor protective potential at the T-cell effector phase. It is possible, in the absence of antigens, that T cells lose this activity over time, which may explain the lack of protection against HHD B-ALL at the T-cell memory phase. Although the T cells induced by EBV LMP1 showed remarkable protection against all tested B-ALL lines, durable protection (complete elimination) was observed only with some of these leukemia lines, presumably due to the higher immunogenicity of those lines.

Differences between human and mouse response to EBV-infected/LMP1-expressing B cells may also inform the interpretation of our findings. The human immune system takes months to completely control a primary EBV infection^17^ in contrast to the mouse immune system where the LMP1^+^ B cells may be cleared within a week^7^. Therefore, it is likely that human TAA-specific T cells may still have tumor-protective potential for some time even after the EBV antibody profile has converted to ‘past infection’ (marked by VCA IgG positivity). These EBV-induced TAA-specific effector T cells, during the acute infection phase, may attack leukemic B cells that express shared TAAs. Moreover, there is a possibility that these T cells also target pre-leukemic B cells for elimination if the latter also express some shared TAAs (see the hypothetical model in Fig. S4A). This protection is particularly applicable to leukemias that have a prenatal origin^22,23^ and might help explain the strong inverse association between HHD B-ALL and EBV seropositivity at ages 2–3. Primary EBV infection at these very early ages would be relatively recent (within the first 2– 3 years of life) and the induced TAA-specific T cells may eradicate pre-leukemic and leukemic HHD B-ALL, which would explain the early protection window seen in the clinical samples. In contrast, if a child contracts EBV at a time when they have no leukemia cells present but have a leukemogenesis later on (postnatal origin), the TAA-specific T cells, now in the memory phase, are ineffective in protection against leukemia development (Fig. S4B). This may explain the lack of association between HHD B-ALL and EBV seropositivity at older ages 4–5 years. Indeed, our finding that the percent of EBV seropositive patients was stable between 2 to 5 years of age (Table 1) supports the possibility of early infection in this cohort. This is also consistent with the previous finding that in US the rate of EBV seropositivity is relatively steady between ages 2–3 and adolescence^10^ (Fig. S5).

Both HHD and *E/R* B-ALL may have pre-natal origins^20,21^,but no association between *E/R* B-ALL and EBV seropositivity was detected at ages 4–5 or ages 2–3. Our model presumes that it is the combination of immunogenicity and timing relative to the primary infection that determine the detectable association. According to this model, HHD B-ALL would be more visible than *E/R* B-ALL to EBV-induced TAA-specific effector T cells, such as through higher numbers and/or expression levels of TAAs. Although this has not been investigated, partly due to the broad and incompletely defined nature of TAAs (including overexpressed antigens, differentiation antigens, and cancer testis antigens)^24^, the finding of a general upregulation of genes/proteins encoded on the gained chromosomes in HHD B-ALL in comparison with *E/R* B-ALL^25^ supports this possibility. There is no suitable HHD B-ALL mouse model for immune function studies. Currently, the only reported HHD B-ALL mouse model arises in *Eμ-ret* mice, driven by a transgene comprising an incidental chimeric human protein (an RFP/RET fusion)^26,27^. The *Eμ-ret* leukemia model has another limitation, as the human protein could be recognized as a xenogeneic antigen and targeted by mouse T cells, particularly in the transplantation setting used in our study.

Reverse causation—that children at ages 2–3 affected by HHD B-ALL cannot develop antibody response to EBV viral infection prior to their leukemia diagnosis—appears unlikely. In most cases, the children’s immune system is likely normal prior to the acute leukemia diagnosis, and indeed some of these children are EBV antibody positive. Additionally, children at these ages affected by other subtypes of B-ALL can mount an antibody response to EBV normally. We also found that children at 4–5 years old affected by HHD B-ALL exhibit clear antibody response to EBV. It is also unlikely that the negative association of HHD B-ALL with EBV is confounded by another co-infecting virus, as high-throughput, large-scale surveys have never detected any virus tending to co-infect humans with EBV^28,29^.

We did not find any B-ALL patients with an EBV antibody profile indicative of acute infection. This might be due to the very short time window during which VCA IgM antibodies are detectable. However, given that our mouse studies demonstrated that LMP1-induced anti-tumor immunity at the effector phase effectively cleared all tested B-cell leukemias, at least transiently, it is also possible that acute EBV infection and B-ALL diagnosis are mutually exclusive. If so, it would suggest potential use of EBV-elicited T cells (manufacturable *in vitro*^19^) to treat B-ALL in patients.

Taken together, our results support the hypothesis that EBV (LMP1)-induced anti-tumor effector T cells may help prevent some EBV-unrelated cancers in early childhood^1^, a notion built upon recent insights into EBV-induced immunity^7-9^. These findings also raise a series of questions for future investigation. First, is HHD B-ALL indeed more immunogenic than *E/R* (and other subtypes) B-ALL because of expressing more TAAs? Second, are TAA-specific memory T cells regulated by immunosuppressive mechanisms^19^ rendering them incapable of leukemia protection, and can they be reinvigorated for leukemia protection? Third, can EBV-induced T cells also help prevent other cancers with high hyperdiploidy in children^30,31^? Fully addressing these questions in future studies may allow us to establish a link between the decrease in the rate of EBV infection in young children and the rise in the rate of childhood cancer in the US^1^ and other developed countries^32^, reappraising the impact of EBV on human cancers.

## MATERIALS AND METHODS

### Mice

C57BL/6J (B6), *Cd19-cre*^*ERT2*^, *Foxp3*^*DTR/GFP*^, and *Eμ-Myc* (all on a B6 background) were obtained from the Jackson Laboratory. *Rag2*^*−/−*^*common γ chain*^*−/−*^ (*Rag2*^*−/−*^*γc*^*−/−*^) mice were bred in-house or purchased from Taconic. The *LMP1*^*flSTOP*^ mice on a B6 background have been described^7^. Homozygous *Cd19-cre*^*ERT2*^ mice were crossed with *LMP1*^*flSTOP*^ heterozygous mice on the B6 background to generate *Cd19-cre* ^*ERT2*^*;LMP1*^*flSTOP*^ (*ERT2-CL*) and *Cd19-cre* ^*ERT2*^ (*ERT2-C*) control mice. To activate Cre^ERT2^, mice were treated with 3 mg (female) or 4 mg (male) tamoxifen (Sigma; dissolved in sunflower oil) by intragastric gavage. *Foxp3*^*DTR/GFP*^;*ERT2-CL* mice were generated by crossing *Cd19-cre*^*ERT2*^*;Foxp3*^*DTR/GFP*^ mice with *LMP1*^*flSTOP*^ mice. Only male *Foxp3*^*DTR/GFP*^;*ERT2-CL* mice were used in experiments (in males, all Foxp3+ CD4+ regulatory T cells express the GFP reporter). Both sexes of all other strains were used (for *in vivo* work, the recipient and donor mice were sex-matched). Mice used in experiments were 2–4 months of age at the start of procedures, except where indicated otherwise. All mice were bred and maintained in the animal facilities at DFCI under specific pathogen-free conditions, with a 12-hour light–dark cycle, ambient temperature of 21–23°C, and humidity of 35–55%. All animal experiments were conducted per protocols approved by the DFCI Institutional Animal Care and Use Committee (IACUC).

### Patient samples

Plasma samples from pediatric B-ALL patients were obtained as part of the DFCI ALL Consortium clinical trials (DFCI 05-001, 11-001, and 16-001). Informed consent was obtained from patients or their legal guardians where applicable. Samples were used only if the patients consented to the future use of the banked samples. The samples were collected at the time of diagnosis, prior to treatment initiation. Peripheral blood was processed by centrifugation to isolate plasma, which was stored at −80°C until use. The samples were de-identified before being released to the investigators for EBV antibody tests. EBV antibody tests were performed in a blinded fashion. Testing was performed on completely de-identified samples and corresponding clinical data, including age at diagnosis and genetic subtype of the disease (based on cytogenetics and FISH reports) was obtained after the EBV serology results. All human sample collection and use were conducted in accordance with protocols approved by the DFCI Institutional Review Board (IRB), in compliance with the Declaration of Helsinki.

### Establishment and culture of leukemia cell lines

Leukemia cells with pre-B cell phenotype (CD19^+^B220^low^IgM^−^c-Kit^−^) were harvested from bone marrow of sick *Eμ-Myc* mice, passaged in wild-type B6 mice, from which the outgrowing leukemia cells were harvested and cultured in media with mouse embryonic fibroblasts (MEF) as feeders until cell lines were established. The culture media were composed of DMEM (Gibco) supplemented with 10% FBS (Sigma), 100 IU/ml penicillin and streptomycin (Gibco), 10 mM HEPES (Corning), 1 mM sodium pyruvate (Gibco), 1X non-essential amino acids (Corning), 50 μM β-mercaptoethanol (Sigma). The established cell lines include L10 (female), L12-3 and L12-33 (female), and L14 (male).

### Generation of *β2m* and *Ciita* knockout mouse leukemia sublines by CRISPR–Cas9

Predesigned CRISPR RNAs (crRNA) targeting *β2m* (5’-CGTGAGTAAACCTGAATCTT-3’) and *Ciita* (5’-AGTCGCTCACTGGTCCCACT-3’ and 5’-CCGTGGACAGTGAATCCACT-3’) were obtained from the Integrated DNA Technologies (IDT). The IDT protocol was followed to prepare the gRNA (crRNA complexed 1:1 with tracrRNA [ATTO 550-labeled]) and RNP complex (formed by gRNA and Alt-R Cas9). The L10 leukemia cell line was transiently electroporated with the RNP (2 μM) supplemented with the Alt-R Cas9 electroporation enhancer (2 μM; IDT) in 100 μL Amaxa P4 solution using Amaxa 4D-Nucleofector system (Lonza) under the program DI-100. Then, cells were transferred into pre-warmed antibiotic-free complete medium and cultured at 37°C. At 48 h post-transfection, ATTO 550-positive (PE^hi^) cells were sorted and expanded in 6-well plate, which were confirmed to be negative for MHC-I or -II expression by flow cytometry.

### Flow cytometry

Single-cell suspensions were stained with monoclonal antibodies specific for mouse B220 (clone RA3-6B2), CD117 (c-kit; clone 2B8), CD4 (clone GK1.5), CD8 (clone 53-6.7), CD19 (clone 6D5), CD69 (clone H1.2F3), IgM (clone RMM-1), Ki67 (clone 16A8), CD279 (PD-1; clone RMP1-30), TCRβ (clone H57-597), IFN-γ (XMG1.2), granzyme B (GzmB; clone NGZB), Foxp3 (clone FJK-16s), Eomes (clone Dan11mag), Tim3 (clone B8.2C12), H2-Kb (clone AF6-88.5), and I-A^b^ (clone AF6-120.1) from BD Biosciences, BioLegend, or eBioscience. Dead cells were excluded using eFluor 506 viability dye (eBioscience). Intracellular staining for GzmB, Foxp3, Eomes, and IFN-γ was performed using the BD Cytofix/Cytoperm staining buffer set (BD Biosciences), according to the manufacturer’s instructions. Samples were acquired on an FACS Fortessa flow cytometer (BD Biosciences) and analyzed using FlowJo software (version 10; Tree Star). FACS sorting was performed using a FACSAria II (BD Biosciences).

### Leukemia models, adoptive T cell transfer, and assessment of leukemia burden

All leukemia models in *ERT2-CL, ERT2-C*, and *Rag2*^*−/−*^*γc*^*−/−*^ mice were established by transplanting 1 x 10^4^ leukemia cells intravenously (i.v., via tail vein). In the case of leukemia model in *Rag2*^*−/−*^*γc*^*−/−*^ mice, three days after leukemia cell transplantation, the mice received, via i.v. injection, 25 x 10^4^ CD4^+^ or CD8^+^ T cells or their combination (25 x 10^4^ cells each) isolated from the spleen of adult *Foxp3*^*DTR/GFP*^;*ERT2-CL* mice. Leukemia burden was assessed by FACS analysis of the peripheral blood sampled at day 21 after leukemia inoculation or collected when mice were euthanized at study endpoints. Red blood cells were lysed using hemolytic Gey’s solution prior to staining. Leukemia cells were detected as CD19^+^B220^low^IgM^−^c-Kit^−^, and the percentage of leukemia cells was calculated relative to total live cells.

### *In vitro* killing assay

Target cells were labelled with CellTrace Violet (Invitrogen) before use. T cells were mixed with labelled target cells at indicated effector-to-target (E:T) cell ratios in 96-well round-bottomed plates. The plates were spun down at 8 g for 2 min before incubation at 37°C for 4 h. Afterward, the cultures were stained for CD4 or CD8 (to exclude effector cells) and active caspase-3 (BD Biosciences), and analyzed by FACS for active caspase-3^+^ CellTrace^+^ cells, which represent apoptotic target cells. The percentage of specific killing = % apoptotic target cells in cultures with both effectors and targets – % apoptotic target cells in cultures with targets alone.

### EBV serology test

Patients’ EBV infection status was determined by testing the presence of IgM and IgG antibodies to EBV viral capsid antigen (VCA) in the plasma samples collected at the time of B-ALL diagnosis. The tests were conducted by the standard approach^7,13^,14 using ELISA kits (Gold Standard Diagnostics), according to the manufacturer’s instructions. The sensitivity and specificity of the kit for VCA IgM were 96.3% and 100%; for VCA IgG 98.0% and 100%, as determined by the manufacturer. Briefly, plasma samples, calibrators, positive and negative controls were pre-diluted per the manufacturer’s instruction before they were dispensed into the test wells (in 96-well plates) and incubated. Afterward, the wells were washed, conjugate was added, followed by another incubation and wash steps. Substrate was added and allowed to incubate for color development before the addition of Stop Reagent. The plates were read at 405 nm in a microplate reader (SpectraMax M3, Molecular Devices LLC) and the optical density (OD) value of each well was recorded, which was then converted into index value according to the formula included in the kit package. Results were interpreted, based on the calculated index value, as positive (≥1.1), negative (<0.9), or equivocal (≥0.9 and <1.1). All samples with positive or negative results (n=172) were included in the data analysis; samples with equivocal results (n=7) were not included in the data analysis. EBV infection status was interpreted as follows: VCA IgM^−^ IgG^−^, no infection; VCA IgM^+^ IgG^+/–^, acute infection; VCA IgM^−^ IgG^+^, past infection.

### Statistical analysis

Mouse survival curves were compared by the log-rank (Mantel-Cox) test; leukemia burden was analyzed by unpaired two-tailed Student’s *t* test, using Prism v.7.03 (GraphPad). EBV serological data were analyzed by Fisher’s exact test. Data are presented as mean ± SD where appropriate, and statistical significance was defined as \**p* < 0.05, ** *p* < 0.01, *** *p* < 0.001, **** *p* < 0.0001; ns, not significant.

## Data Availability

All data produced in the present study are available upon reasonable request to the authors

## Acknowledgments

We thank the DFCI Hematologic Neoplasia Flow Cytometry Core for excellent assistance with the flow cytometry studies and cell sorting, R. Avalos and D. McDermott for administrative assistance. This work was supported by DFCI Lymphoma Research Center Grant to B.Z.; Claudia Adams Barr Program for Innovative Cancer Research Award to I.-K.C.

## Author contributions

J.P. and I.-K.C planned and conducted the mouse studies with Z.W. and J.G.; Z.W. performed the EBV serological study; C.S., S.R., V.K., and M.H.H. retrieved the plasma samples and correlative data used in the study; Y.F. contributed to study design and analysis of data derived from B-ALL patient samples; J.R. provided scientific advice; M.S. advised on study design; L.V., A.E.P., M.B., and L.B.S. contributed patient samples and correlative data; Y.P. coordinated with B.Z. on study design, provided patient samples and correlative data, and contributed in data interpretation; B.Z. conceived and supervised the overall research; J.P., Y.P. and B.Z. wrote the manuscript, with input from all authors.

## REFERENCES

1. Zhang, B., Choi, I.K., Panaampon, J. & Wang, Z. Does delayed EBV infection contribute to rising childhood cancers? Trends Immunol 43, 956–958 (2022).

2. Greaves, M. A causal mechanism for childhood acute lymphoblastic leukaemia. Nature reviews. Cancer 18, 471–484 (2018).

3. Gilham, C., et al. Day care in infancy and risk of childhood acute lymphoblastic leukaemia: findings from UK case-control study. BMJ 330, 1294 (2005).

4. Hauer, J., Fischer, U. & Borkhardt, A. Toward prevention of childhood ALL by early-life immune training. Blood 138, 1412–1428 (2021).

5. Cobaleda, C., Vicente-Duenas, C. & Sanchez-Garcia, I. An immune window of opportunity to prevent childhood B cell leukemia. Trends Immunol 42, 371–374 (2021).

6. Cobaleda, C., Vicente-Duenas, C. & Sanchez-Garcia, I. Infectious triggers and novel therapeutic opportunities in childhood B cell leukaemia. Nature reviews. Immunology 21, 570–581 (2021).

7. Choi, I.K., et al. Mechanism of EBV inducing anti-tumour immunity and its therapeutic use. Nature 590, 157–162 (2021).

8. Long, H.M., et al. CD4+ T-cell clones recognizing human lymphoma-associated antigens: generation by in vitro stimulation with autologous Epstein-Barr virus-transformed B cells. Blood 114, 807–815 (2009).

9. Linnerbauer, S., et al. Virus and autoantigen-specific CD4+ T cells are key effectors in a SCID mouse model of EBV-associated post-transplant lymphoproliferative disorders. PLoS pathogens 10, e1004068 (2014).

10. Hjalgrim, H., Friborg, J. & Melbye, M. The epidemiology of EBV and its association with malignant disease. in Human Herpesviruses: Biology, Therapy, and Immunoprophylaxis (eds. Arvin, A., et al.) (Cambridge, 2007).

11. Vrooman, L.M., et al. Refining risk classification in childhood B acute lymphoblastic leukemia: results of DFCI ALL Consortium Protocol 05-001. Blood Adv 2, 1449–1458 (2018).

12. Place, A.E., et al. Intravenous pegylated asparaginase versus intramuscular native Escherichia coli L-asparaginase in newly diagnosed childhood acute lymphoblastic leukaemia (DFCI 05-001): a randomised, open-label phase 3 trial. Lancet Oncol 16, 1677–1690 (2015).

13. Balfour, H.H., Jr., et al. Age-specific prevalence of Epstein-Barr virus infection among individuals aged 6-19 years in the United States and factors affecting its acquisition. The Journal of infectious diseases 208, 1286–1293 (2013).

14. Dowd, J.B., Palermo, T., Brite, J., McDade, T.W. & Aiello, A. Seroprevalence of Epstein-Barr virus infection in U.S. children ages 6-19, 2003-2010. PloS one 8, e64921 (2013).

15. Gu, Z., et al. PAX5-driven subtypes of B-progenitor acute lymphoblastic leukemia. Nature genetics 51, 296–307 (2019).

16. Moorman, A.V. The clinical relevance of chromosomal and genomic abnormalities in B-cell precursor acute lymphoblastic leukaemia. Blood Rev 26, 123–135 (2012).

17. Odumade, O.A., Hogquist, K.A. & Balfour, H.H., Jr. Progress and problems in understanding and managing primary Epstein-Barr virus infections. Clin Microbiol Rev 24, 193–209 (2011).

18. CDC. Laboratory Testing for Epstein-Barr Virus (EBV). (2024).

19. Zhang, B. & Choi, I.K. Facts and Hopes in the Relationship of EBV with Cancer Immunity and Immunotherapy. Clinical cancer research : an official journal of the American Association for Cancer Research 28, 4363–4369 (2022).

20. Yasuda, T., et al. Studying Epstein-Barr virus pathologies and immune surveillance by reconstructing EBV infection in mice. Cold Spring Harbor symposia on quantitative biology 78, 259–263 (2013).

21. Harris, A.W., et al. The E mu-myc transgenic mouse. A model for high-incidence spontaneous lymphoma and leukemia of early B cells. The Journal of experimental medicine 167, 353–371 (1988).

22. Hein, D., Borkhardt, A. & Fischer, U. Insights into the prenatal origin of childhood acute lymphoblastic leukemia. Cancer Metastasis Rev 39, 161–171 (2020).

23. Marcotte, E.L., Spector, L.G., Mendes-de-Almeida, D.P. & Nelson, H.H. The Prenatal Origin of Childhood Leukemia: Potential Applications for Epidemiology and Newborn Screening. Front Pediatr 9, 639479 (2021).

24. Gires, O. & Seliger, B. Tumor-Associated Antigens: Identification, Characterization, and Clinical Applications, Part one, 1–43 (2009).

25. Yang, M., et al. Proteogenomics and Hi-C reveal transcriptional dysregulation in high hyperdiploid childhood acute lymphoblastic leukemia. Nat Commun 10, 1519 (2019).

26. Farrokhi, A., et al. The Emu-Ret mouse is a novel model of hyperdiploid B-cell acute lymphoblastic leukemia. Leukemia 38, 969–980 (2024).

27. Wasserman, R., Zeng, X.X. & Hardy, R.R. The evolution of B precursor leukemia in the Emu-ret mouse. Blood 92, 273–282 (1998).

28. Xu, G.J., et al. Viral immunology. Comprehensive serological profiling of human populations using a synthetic human virome. Science 348, aaa0698 (2015).

29. Kamitaki, N., Tang, D., McCarroll, S.A. & Loh, P.R. The DNA virome varies with human genes and environments. Nature (2026).

30. Look, A.T., Hayes, F.A., Nitschke, R., McWilliams, N.B. & Green, A.A. Cellular DNA content as a predictor of response to chemotherapy in infants with unresectable neuroblastoma. The New England journal of medicine 311, 231–235 (1984).

31. Douglass, E.C., et al. Hyperdiploidy and chromosomal rearrangements define the anaplastic variant of Wilms’ tumor. Journal of clinical oncology : official journal of the American Society of Clinical Oncology 4, 975–981 (1986).

32. Steliarova-Foucher, E., et al. Changing geographical patterns and trends in cancer incidence in children and adolescents in Europe, 1991-2010 (Automated Childhood Cancer Information System): a population-based study. Lancet Oncol 19, 1159–1169 (2018).

