## Supplemental Information for "Exclusion of the commonest subtype of B-cell leukemia in children acquiring EBV infection in early life"

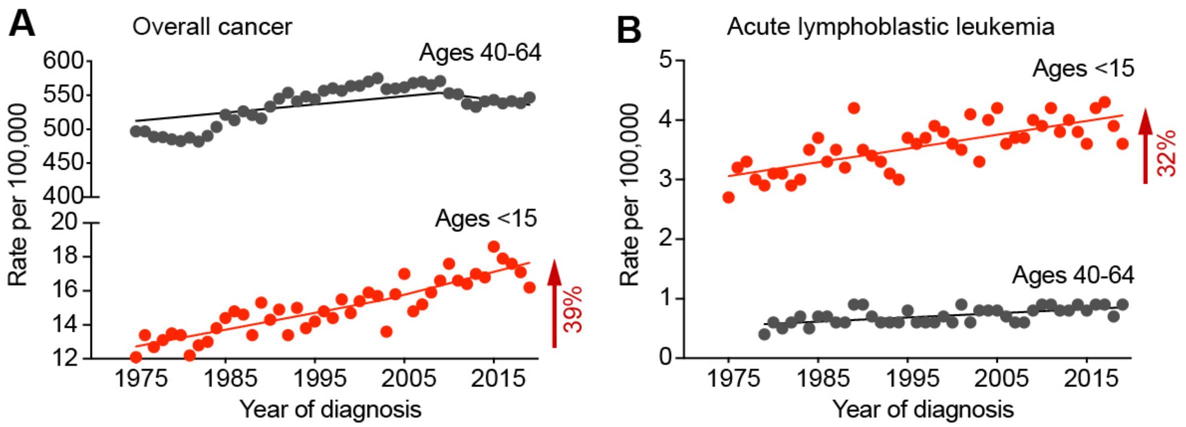


**Figure S1. The rise of childhood cancers in the US.** Trends in overall cancer rates (**A**) and acute lymphoblastic leukemia (ALL) rates (**B**) in US populations aged <15 vs 40-64. Most (~85%) of childhood ALL are of the B-cell type (B-ALL). Graphs are based on the National Cancer Institute (NCI) Surveillance, Epidemiology, and End Results (SEER) data from 1975-2019.

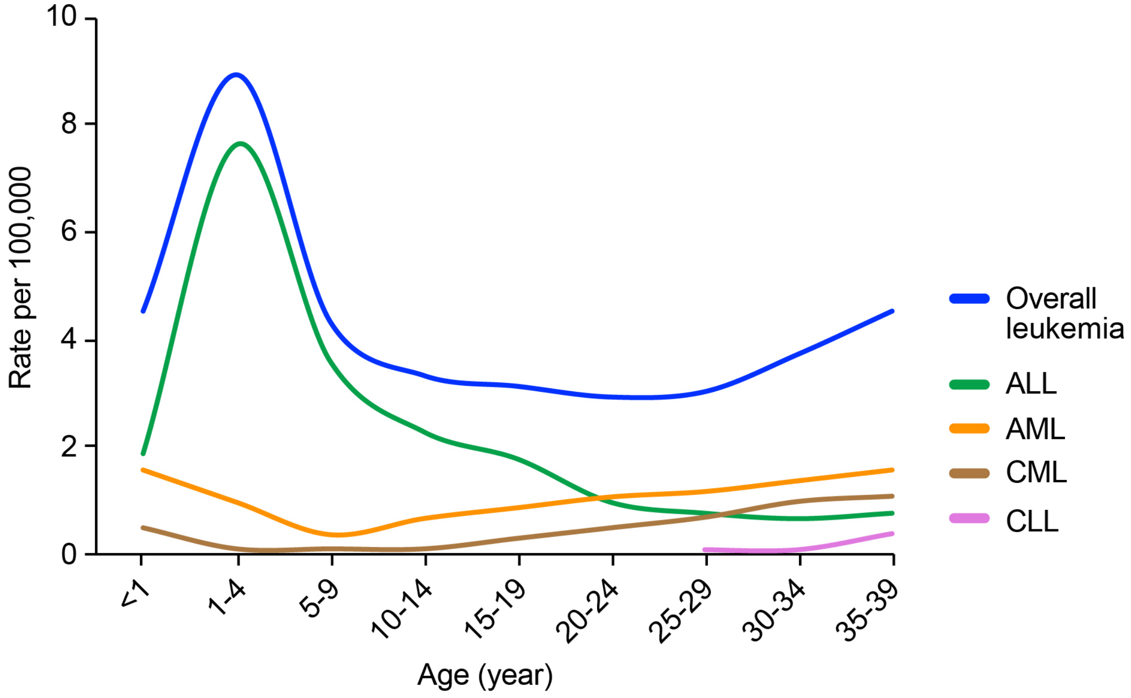


**Figure S2. Incidence rates of ALL and other leukemias in US populations at the indicated ages.** AML, acute myeloid leukemia; CML, chronic myeloid leukemia; CLL, chronic lymphocytic leukemia. Graphs are based on the NCI SEER data 2012-2016.

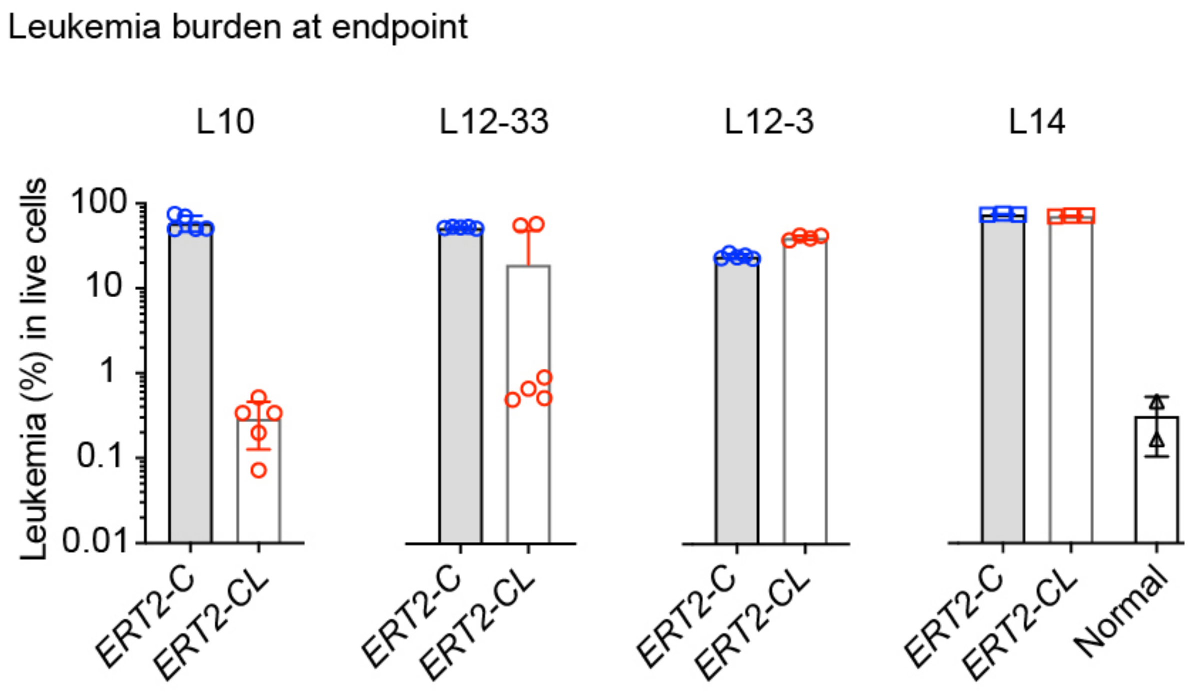


**Figure S3. Leukemia burden in peripheral blood of *ERT2-C* and *ERT2-CL* mice at the end point shown in Fig. 1B.** Leukemia ($CD19^{+}B220^{low}IgM^{-}c\text{-}Kit^{-}$) burden was assessed by FACS; Normal, untreated wild-type mice as control to indicate background staining. Data are presented as mean ± SD.

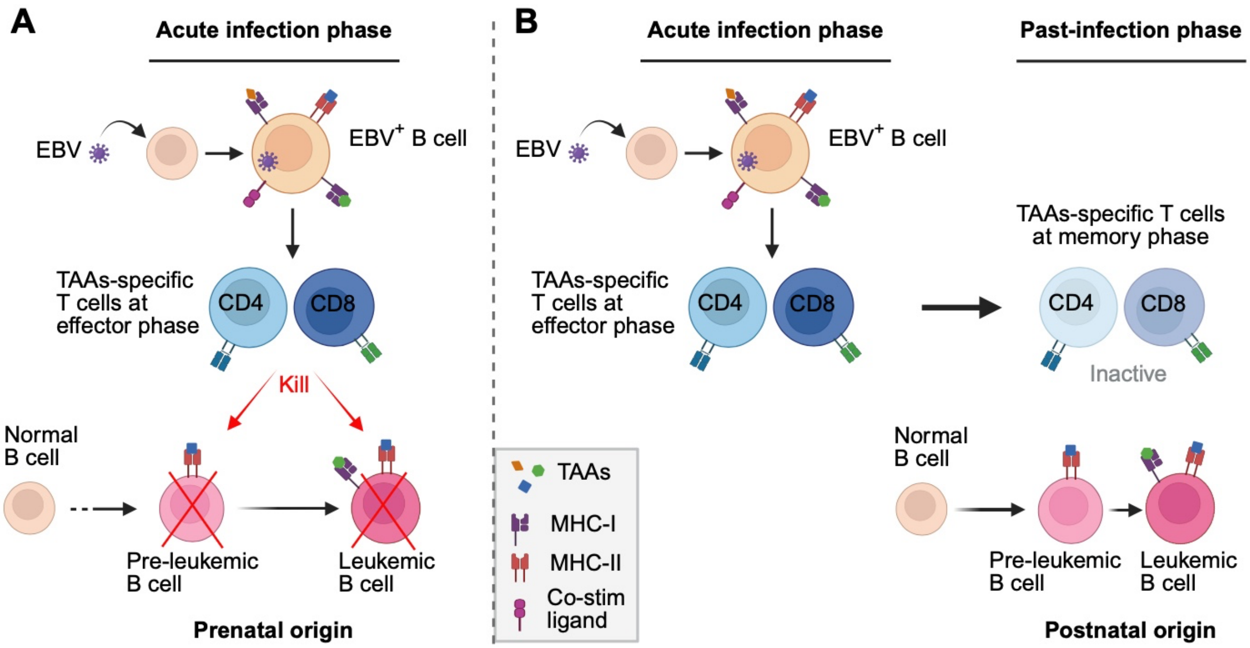


**Figure S4. A hypothetical model depicting scenarios where EBV-induced TAA-specific T cells may be capable or incapable of protecting against pediatric B-ALL. A**, At 'acute infection phase', TAA-specific T cells elicited by EBV-infected/-transformed B cells are functionally active (at effector phase) and able to recognize and kill existing leukemic/pre-leukemic B cells that express shared TAAs. **B**, At 'past-infection phase', the TAA-specific T cells (now at memory phase) become dull/inactive and unable to kill leukemia developing at this time. See text for further details.

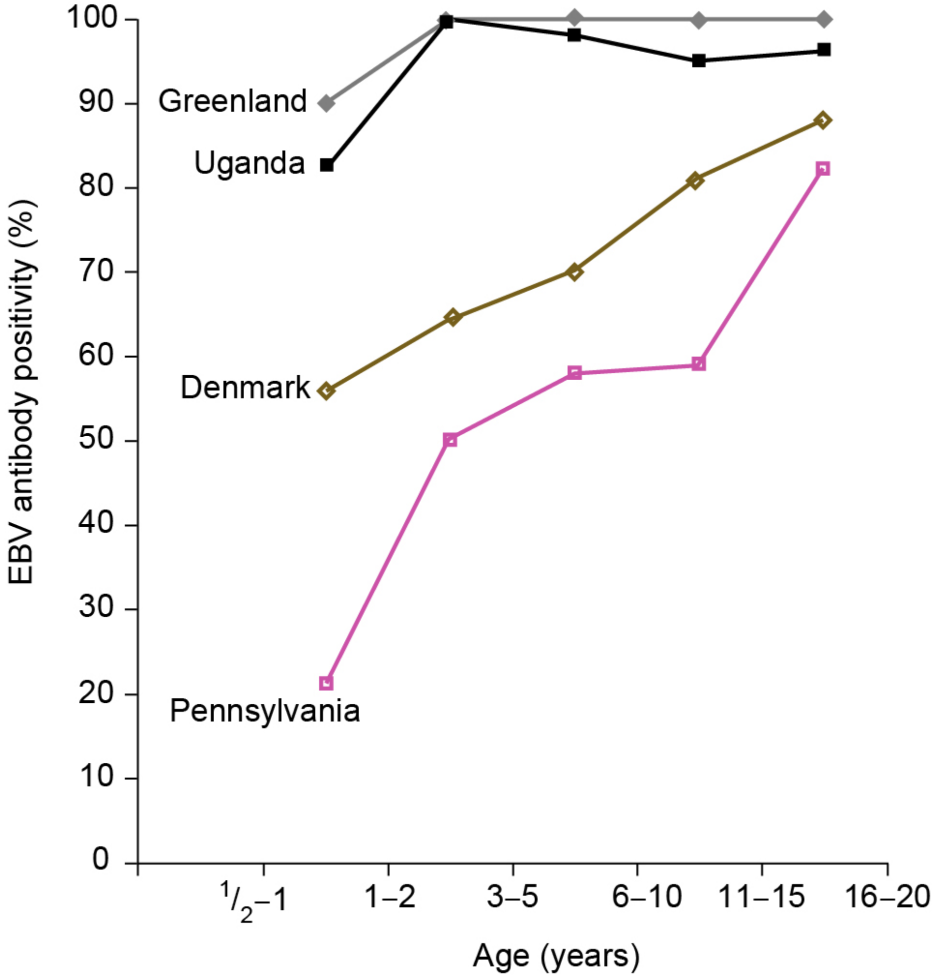


**Figure S5. EBV seroprevalence in populations of the indicated countries/regions.** Graph drawn based on reference 10 (data for children < 1-year old are not shown, because EBV antibody test at this age may be unreliable due to potential presence of maternal antibodies).
